# The LMSz method - an automatable scalable approach to constructing gene-specific growth charts in rare disorders

**DOI:** 10.1101/2024.08.19.24312213

**Authors:** K.J. Low, J. Foreman, R.J. Hobson, H. Kwuo, E. Martinez-Cayuelas, B. Almoguera, P. Marin-Reina, S.G. Caraffi, L. Garavelli, E. Woods, M. Balasubramanian, A. Bayat, C.W. Ockeloen, C.M. Wright, H.V. Firth, T.J. Cole

## Abstract

**Introduction:** Children with monogenic neurodevelopmental disorders often grow abnormally. Gene-specific growth charts would be useful but require large samples to construct them using the conventional LMS method.

**Methods:** We transformed anthropometry to British 1990 reference z-scores for 328 UK and 264 international probands with *ANKRD11, ARID1B, ASXL3, DDX3X, KMT2A* or *SATB2-*related disorders, and modelled mean and standard deviation (SD) of the z-scores as gene-specific linear age trends adjusted for sex. Back-transforming the mean ±2 SD lines provided gene-specific median, 2^nd^ and 98^th^ centiles.

**Results:** The resulting z-score charts look plausible on several counts. Only *KMT2A* shows a (rising) age trend in median height, while BMI and weight increase in several genes, possibly reflecting population trends. Apart from *SATB2* and *DDX3X,* the gene-specific medians are all below the reference (range 0.1^th^ centile for height *KMT2A* to 36^th^ centile for BMI *ANKRD11*). Median OFC shows no age trend, with medians ranging from 10^th^-30^th^ centile, and *ASXL3* lowest, on the 3^rd^ centile. There are no sex differences in 19/24 cases.

**Conclusions:** Our LMSz method produces gene-specific growth charts for rare diseases, an essential clinical tool for paediatric care. We plan to automate it within the DECIPHER platform, enabling availability for all relevant genes.

## Introduction

Health professionals measure and plot children’s growth at intervals on appropriate charts to track the trajectory against reference centiles (1). Many children with genetic disorders plot on an extreme centile—in itself a clue to a possible underlying genetic diagnosis (2). Once a genetic diagnosis is made, plotting on a standard chart may be misleading. It may, for example, suggest a child is of short stature and underweight where they are growing normally for their genetic disorder leading to clinical/parental anxiety resulting in unnecessary investigation and unwarranted/ineffective intervention. Conversely, abnormal growth may be incorrectly ascribed to the underlying genetic disorder, and other causes left untreated. Gene-specific growth charts are therefore important in paediatric care. However, due to the small numbers of affected individuals, few gene-specific growth charts are available (3–5).

This proof-of-principle study tests a new method for constructing growth charts in rare disorders based on small datasets, which we call the LMSz method. We call it this because it applies the LMS method (9) on the z-score scale.

## Methods

Growth data were collated from the Deciphering Developmental Disorders (DDD) (6) and GenROC study (7) datasets, both from the UK. For proof-of-principle we selected *ANKRD11*, a frequently diagnosed gene in the DDD study with a well described phenotype. We included only participants with pathogenic or likely pathogenic *ANKRD11* variants, excluding any with composite genetic diagnoses. We extracted data on sex, gestational age, birth weight, birth occipitofrontal circumference (OFC), and longitudinal measures of height, weight and OFC and the associated ages at measurement.

For *ANKRD11*, we undertook a second phase of analysis including European datasets (Spain, Denmark and The Netherlands). We also extended the analysis to five other genes: *ARID1B*, *ASXL3*, *DDX3X*, *KMT2A* and *SATB2*. *ASXL3* growth data were collated from the DDD study and the *ASXL3* international natural history study (IRAS: 316055). We compared our *ARID1B* charts with the Coffin Siris Syndrome heterogeneous gene group growth charts (4).

We also analysed data from the Mowat Wilson Syndrome (MWS) Growth Chart Consortium and compared our results with those obtained by fitting the raw data using the LMS method (see Supplement 1 for details). They were also compared to the corresponding centile charts published by the MWS Consortium (3).

Age was adjusted for gestation using the formula *adjusted age* = *age* + (*gestation* - 40) × 7/365.25. Body mass index (BMI) was calculated as *weight*/*height*^2^ in units of *kg*/*m*^2^.

### Statistical analysis

Growth centile charts are conventionally constructed using the LMS (lambda-mu-sigma) method (8), a special case of the family of Generalized Additive Models for Location Scale and Shape (GAMLSS) (9). Using anthropometry from a reference sample of children the LMS method estimates three smooth curves in age. M is the median or 50^th^ centile curve; the S curve is the coefficient of variation or fractional standard deviation (SD), and the L curve is a measure of skewness. Any required centile curve can be constructed from the LMS curves, and by reversing the process data for individuals can be converted to exact z-scores.

The LMS method is most effective with large datasets, ideally many thousands of individuals (10). The issue with charts for rare diseases is the inherent scarcity of data. In this case, the chart cannot be based solely on the available data, it must “borrow strength” from previous knowledge about how children grow, as contained in an existing or “baseline” growth reference. This baseline reference can then be adjusted using the rare disease data to obtain a reference for the rare disease. This is analogous to a Bayesian analysis where prior information (the baseline reference) is combined with rare disease data to give a posterior or updated reference relevant to the rare disease.

The process is as follows: the data are first converted to z-scores using the baseline reference, which here is the British 1990 reference (UK90) (11), the official UK chart for birth and age 4-20 and the reference used by the DECIPHER database. Switching to the z-score scale creates centile curves which are horizontal lines, as each centile curve connects points with the same z-score; they are also the same for both sexes. For representative children the z-transformed centiles are Normally distributed with mean (and median) 0 and SD 1 at all ages. Both mean and SD can then be viewed as straight lines plotted against age. Other centile curves are lines above or below the median, spaced according to the corresponding z-score (e.g. the 97.7^th^ centile is at z = 2, i.e. 2 SDs above the mean).

The next stage is to assume that the rare disease median curve is also a straight line but estimated from the rare disease data as the linear regression of UK90 z-score on age and sex. The SD curve is estimated in the same way from the linear regression of the z-score SD on age and sex. GAMLSS is used to estimate the mean and SD regression lines simultaneously, using the Normal or NO distribution family. The two lines are analogous to the M and S curves of the LMS method, but on the z-score scale. Note that there is no equivalent L line as the normal distribution is by definition not skew, and the conversion from measurement to z-score is assumed to have removed any skewness.

The GAMLSS regression equation for the M line is made up of four terms: intercept, age trend, mean sex difference and age by sex interaction (i.e. the age trend differing between the sexes), and the coefficient for each term is estimated:

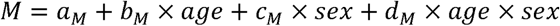

But if one or more of the fitted coefficients is small enough, the corresponding term(s) can be dropped from the regression, leading to a set of five progressively simpler equations:

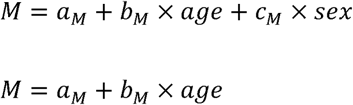

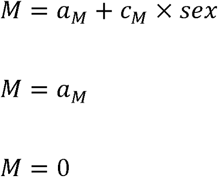

The SD or S line is modelled in the same way, except it is log transformed to ensure positivity. The interaction term is also omitted for simplicity.

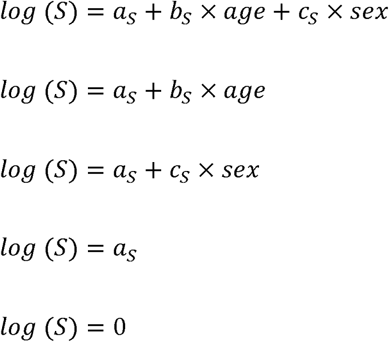

Note that the final equations in these lists are null models where the coefficient is 0. They correspond to M = 0 and S = 1, the mean and SD for the baseline UK90 reference, which are also the default values for the GAMLSS NO distribution. GAMLSS estimates the M line and S line simultaneously, and there are 6 M line equations and 5 S line equations, so there are 6x 5 = 30 possible model combinations. All 30 are fitted to the data in turn, and they are compared for goodness of fit using either the Akaike information criterion (AIC) or the Bayesian information criterion (BIC), the optimal model being the one with the lowest AIC or BIC. Note that the S line is linear on the log scale, but backtransformed it is slightly curved if the *b*_S_ coefficient is non-zero.

Lack of fit is measured by the deviance, or -2 log likelihood. The AIC penalises the deviance by adding two units for each coefficient in the model, while the BIC uses a larger penalty of log *n* units per coefficient, where *n* is the sample size. This penalises more complex models, which tend to fit better.

For a model where *n* = 7 the AIC and BIC are the same, but for larger *n* the BIC penalty is larger. For example, the most numerous measurement is weight in *ANKRD11* where there are 488 points and *log* (488) ≈ 6, so here the BIC penalty is three times the AIC penalty. For this reason, the BIC models are either the same as or simpler than the AIC models.

For larger samples the set of models for the M line can be extended to include a cubic P-spline curve in age as an alternative to a straight line. This increases the number of possible fitted M lines from 6 to 9, and the total of combined M+S models from 30 to 45.

Once the optimal model has been identified, it is used to predict the required centile lines on the UK90 z-score scale. They are then backtransformed to the measurement scale using the UK90 reference, giving a set of centile curves appropriate for the rare disease.

### Error-checking

To detect possible data outliers, the data in the form of UK90 z-scores were centred and scaled to gene-specific z-scores, and z-scores exceeding 3.5 in absolute value were excluded from the analysis. In this way several data errors were identified and either corrected or excluded.

For simplicity the analysis treats any repeated measures in the data as independent.

### Specificity

To test the specificity of the method, z-scores of measurements with *ANKRD11* (being the most numerous) were replaced by z-scores randomly sampled from a standard Normal distribution, with mean 0 and SD 1. In this way simulated z-score data were generated with the age and sex structure of the underlying data, where the optimal model ought to be *M* =0 and *log* (*S*) = 0, corresponding to the baseline UK90 reference. The model was then fitted repeatedly to newly sampled data, and the optimal model each time noted, 100 times each for height (n = 355 points), weight (n = 488), BMI (n = 343) and OFC (n = 231).

## Results

Table 1 provides counts of the number of subjects and data by gene, country, sex and measurement. The age range was restricted to 0-18 years. Linked ethnicity data were not recorded, but we assume the vast majority of cases were Caucasian based on the sources.

**Table 1:** counts of the number of subjects and data by gene, country, sex and measurement. *NatS = Natural History Study (International)

| Gene | Source | Individuals |  | Height |  | Weight |  | BMI |  | OFC |  | Total |
| --- | --- | --- | --- | --- | --- | --- | --- | --- | --- | --- | --- | --- |
|  |  | Female | Male | Female | Male | Female | Male | Female | Male | Female | Male |  |
| ANKRD11 | DDD | 34 | 50 | 31 | 46 | 60 | 95 | 28 | 43 | 36 | 53 | 392 |
| ANKRD11 | GenROC | 5 | 6 | 6 | 11 | 8 | 18 | 5 | 10 | 2 | 6 | 66 |
| ANKRD11 | Spanish | 33 | 35 | 54 | 78 | 58 | 88 | 53 | 76 | 33 | 48 | 488 |
| ANKRD11 | Danish | 3 | 6 | 13 | 13 | 13 | 14 | 13 | 13 | 6 | 3 | 88 |
| ANKRD11 | Dutch | 26 | 40 | 45 | 58 | 64 | 70 | 45 | 57 | 20 | 24 | 383 |
| ARID1B | DDD | 38 | 34 | 33 | 24 | 70 | 58 | 29 | 22 | 41 | 33 | 310 |
| ASXL3 | DDD/NatS* | 49 | 70 | 36 | 52 | 83 | 118 | 32 | 48 | 24 | 41 | 434 |
| DDX3X | DDD | 48 | 5 | 36 | 4 | 84 | 8 | 34 | 3 | 47 | 6 | 222 |
| KMT2A | DDD | 29 | 43 | 21 | 37 | 50 | 76 | 21 | 32 | 22 | 44 | 303 |
| KMT2A | GenROC | 1 | 4 | 3 | 13 | 4 | 18 | 3 | 13 | 1 | 11 | 66 |
| SATB2 | DDD | 12 | 21 | 9 | 15 | 20 | 37 | 7 | 13 | 12 | 24 | 137 |
| Total |  | 278 | 314 | 287 | 351 | 514 | 600 | 270 | 330 | 244 | 293 | 2889 |

### Z-score centile charts and growth charts

The results are presented as a rectangular array of centile charts by gene (*ANKRD11, ARID1B, ASXL3, DDX3X, KMT2A* and *SATB2*) for height, weight, BMI, and OFC (Figure 1). Three centiles are displayed, corresponding to z-scores -2, 0 and 2, i.e. the 2.3^rd^, 50^th^ and 97.7^th^ centiles. For simplicity they are referred to here as the 2^nd^, 50^th^ and 98^th^ centiles.

**Figure 1:**
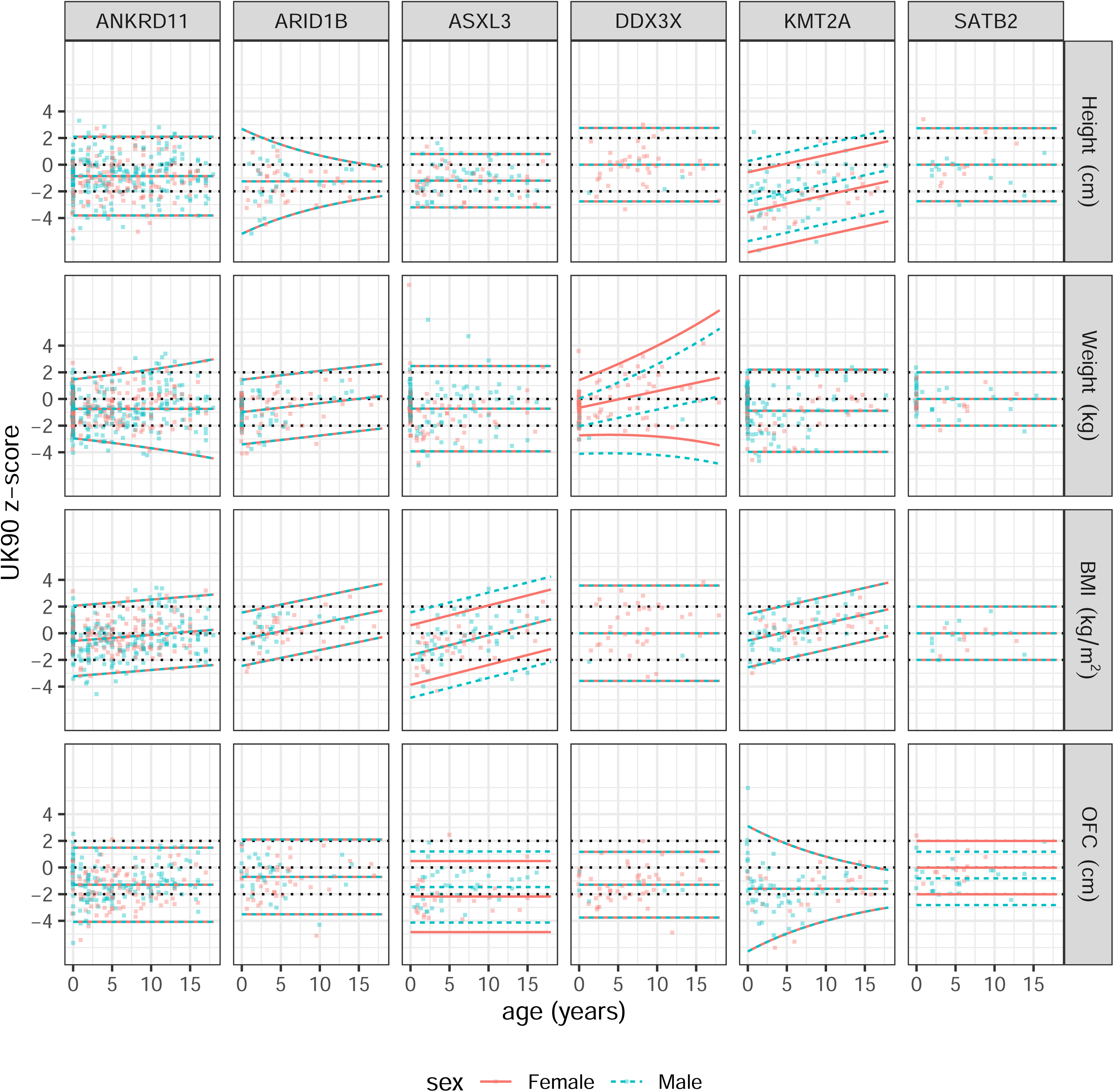
mean ±2 SD lines representing the 2^nd^, 50^th^ and 98^th^ gene-specific z-score “centiles” for height, weight, BMI and OFC across the genes *ANKRD11, ARID1B, ASXL3, DDX3X, KMT2A and SATB2*. The corresponding UK90 z-score centiles at -2, 0 and 2 are shown as dashed lines.

In the five plots where there is a sex difference the centile lines are coloured red for females and turquoise for males. The raw data are also shown, colour coded similarly. The *ANKRD11* charts use the data from all four countries, while Supplement 2 discusses the appreciable inter-country differences in *ANKRD11* height.

Figure 1 penalises the plots using the BIC, while Supplementary Figure 2 shows the same plots penalised with the AIC. As expected, the two sets of centiles are broadly similar, though 13 of the 24 plots in SF2 show a sex difference as against five in Figure 1. Similarly, Supplementary Figure 3 shows the BIC plots of Figure 1 with the option of a spline curve for the M line. Five of the 24 facets select a spline curve, and in all cases they show a dip in early life compared to later childhood.

The specificity of the method was assessed with the *ANKRD11* data. Using the BIC models, respectively 98, 97, 97 and 97 times out of 100 the selected model was the null model, i.e. appropriate for the baseline growth reference. This corresponds to an average specificity of just over 97%, i.e. the percentage of times the procedure correctly chooses the baseline reference for data from non-syndromic children. For comparison, repeating the exercise using the AIC models gave a much lower specificity of 53%.

Overall, the differences between Figure 1 and Supplementary Figures 2 and 3 are small, and given their lower specificity there is no obvious reason to prefer the more complicated AIC centiles over the BIC-based centiles, so the focus from here on is the BIC centiles of Figure 1.

Supplementary Table 1 shows the regression coefficients and standard errors for the models in Figure 1. Except for weight and BMI for *SATB2*, all the models include one or more significant coefficients, some of them highly so, despite the small sample sizes involved. Conversely no model includes an age by sex interaction even though it was an option. Altogether there are 53 regression coefficients fitted, which with 24 distinct gene-measure models correspond to just over two coefficients per model. Supplementary Table 2 shows the 74 corresponding coefficients for the AIC models.

Figures 2 and 3 show the 2^nd^, 50^th^ and 98^th^ measurement centiles obtained by back-transforming the z-score centiles of Figure 1, for females and males respectively, as a rectangular 4 x 6 plot by measurement and gene, with the corresponding UK90 centiles shown as dashed lines.

**Figure 2:**
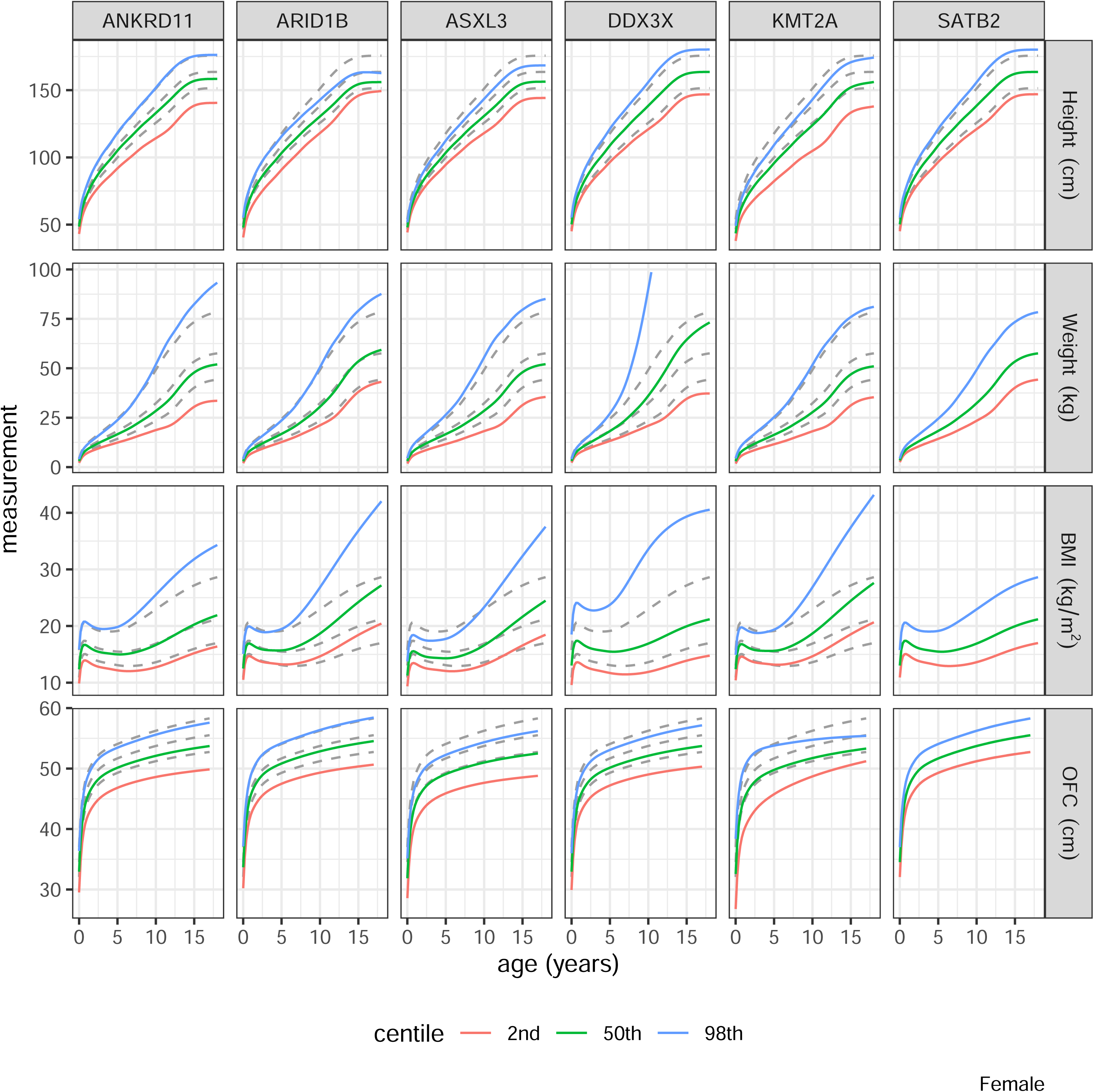
measurement centiles for females by measurement and gene obtained by back-transforming the z-score centiles shown in Figure 1.

**Figure 3:**
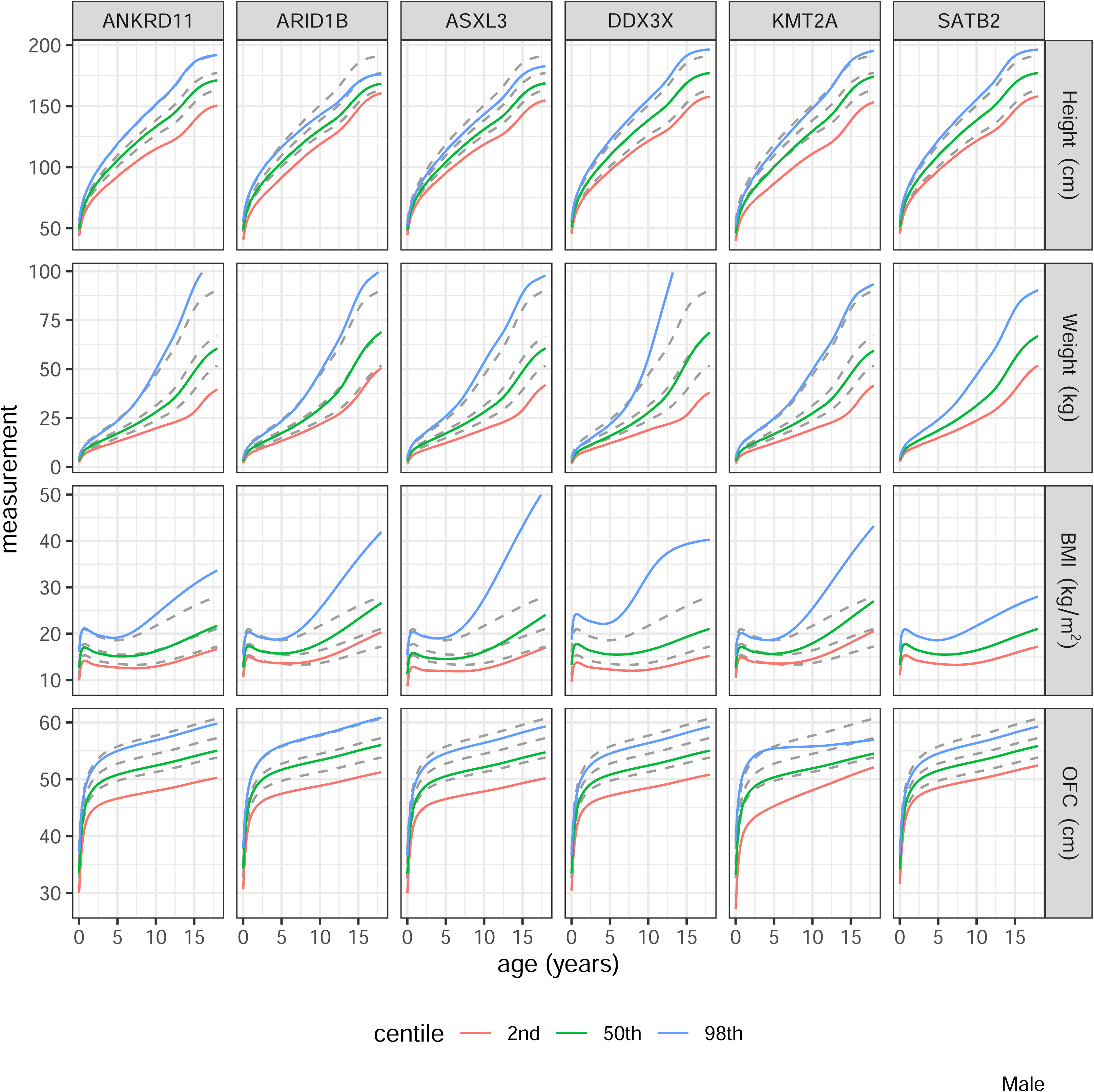
measurement centiles for males by measurement and gene obtained by back-transforming the z-score centiles shown in Figure 1.

### Gene-specific charts

In 19 of the 24 panels in Figure 1, the centiles for males and females are the same, including all four measurements for *ANKRD11*, *ARID1B* and *KMT2A*. The sex differences where they appear are generally small. None of the genes show an age trend in median OFC, and only *KMT2A* shows a (rising) age trend in median height. But for weight and BMI two and four genes respectively show rising trends, by up to +2 SDs from birth to age 18. This may reflect increasing adiposity with age, as the UK90 reference corresponds to child adiposity as seen in 1990, and obesity prevalence increases with age (12). In 20 of the panels the three centiles are parallel lines, indicating that the variability is constant across age. The most striking exception is for weight with *DDX3X*, where the variability is dramatically greater at age 18 than at birth.

Supplementary Table 3 shows the UK90 centiles at birth corresponding to the 2^nd^, 50^th^ and 98^th^ centiles for each measurement, averaged across sex, by gene.

### 50^th^ centile

The median for SATB2 (height, weight and BMI) and for DDX3X (height and BMI) correspond to the UK90 median. However for all other measurements and genes the median is much lower than reference, ranging from 0.08th (height KMT2A) to 28^th^ (BMI ANKRD11) UK90 centile.

### 98th centile

The 98^th^ centiles are nearly all above the UK90 90^th^ centile. However 98^th^ centile for height in *KMT2A* is on the 44^th^ UK90 centile. Very few *KMT2A* cases achieve even median height on the UK90 chart. Other relatively low 98^th^ centiles are for height in *ASXL3* (79^th^) and weight in *DDX3X* (76^th^).

### 2nd centile as z-score

The 2^nd^ centiles at birth are shown as UK90 z-scores because many are far below the 1^st^ UK90 centile. The most extreme cases are *KMT2A* (-6 SD) and *ARID1B* (-5 SD), while all the 2^nd^ centiles are on or below the UK90 2^nd^ centile.

### Mowat Wilson Syndrome (MWS) – ZEB2

Given the large sample size, we estimated MWS centiles in two ways: the LMSz method as applied to the z-scores (Figure 4A), and the LMS method as applied to the raw data (Figure 4B). On the z-score scale (Figure 4A) all four measurement medians are below the UK90 median at birth, with the OFC median close to the UK90 2^nd^ centile. The medians all fall further with increasing age, and at faster rates for weight and OFC among the females. Figure 4B compares the 2^nd^, 50^th^ and 98^th^ centiles by the two methods, for the sexes separately, where the solid curves correspond to the LMS method while the dashed curves are for the LMSz method. For height, weight and BMI there is reasonable agreement, though the BMI 98^th^ centiles agree less well. The height and weight centiles are consistent by sex, being similar until puberty when the males become taller and heavier. However, unlike the LMSz centiles, the LMS centiles continue rising after puberty despite growth ending, because there are too few data at older ages to pull them down. OFC performs poorly, with all three LMS-based centiles rising and falling at different ages, while the LMSz centiles rise linearly for males but fall in later childhood for females.

**Figure 4:**
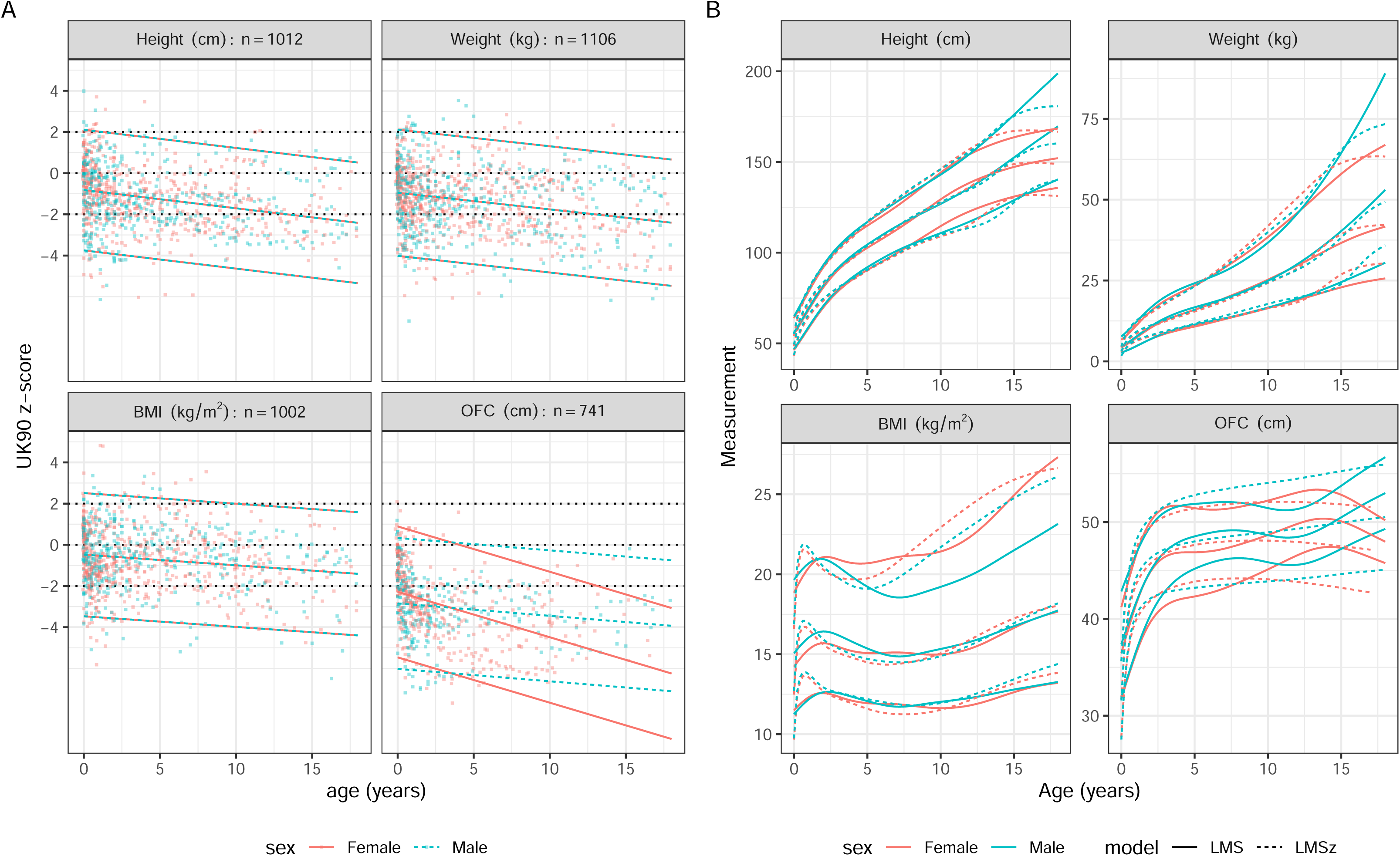
A) MWS data on the z-score scale, in the same form as Figure 1. B) Comparison of the 2^nd^, 50^th^ and 98^th^ centiles by the two methods, for the sexes separately - solid curves correspond to the LMS method and dashed curves to the LMSz method.

## Discussion

We have developed a method for producing gene-specific z-score centile charts and growth charts based on small datasets in rare disease. The resulting charts are unbiased due to the way they are constructed, cosmetically they look plausible, they agree with other published literature, and the centiles almost always exactly match the baseline reference when given random z scores ie. the specificity is as high as 97%.

### Sample size

Growth charts typically require many thousands of data points to ensure adequate precision for the outer centiles (10). This does not work for the small samples available in rare disease cohorts. The LMSz method instead considers the universe of all possible simple models, such as all combinations of sex and linear age for the mean and SD, and selects the best model, penalised for complexity. This works because the universe of all possible linear models is small, between 30 and 45 alternative models, and the model selection does not depend on statistical significance.

### Fitting of centiles

Due to the larger penalty per extra df, the BIC models ought to be simpler than the AIC models, and indeed they are—the BIC models underlying the 24 facets in Figure 1 involve 53 coefficients, i.e. ∼2 coefficients per facet. In contrast the corresponding AIC models in Supplementary Figure 2 involve 74 coefficients, averaging over 3 per facet. Despite this the centiles in Figure 1 and Supplementary Figure 2 are not materially different, probably because the coefficients are all small. The specificity calculation showed that the AIC models had only 53% specificity, compared to 97% for the BIC models, for selecting the correct null model when the data were random Normal. This shows that AIC tends to choose over-complex models, which in turn supports the use of the BIC models. Note that the sensitivity of the method cannot be tested in the same way as the specificity. It involves knowing in advance the centiles appropriate for the particular genotype, which are by definition unknown.

### Z-scores versus growth centiles

The z-score centile plots show clearly the differences in growth pattern for a particular gene compared to UK90. The z-score plots compactly show growth trends over time, and sex differences where present. However, in clinical practice health professionals measure and plot their patients’ growth on centile charts as part of standard clinical care. For this reason, it is important to provide the back-transformed centile charts for each gene, as in Figure 2 and 3, as clinicians will find them more familiar in measurement units.

### Comparison with the literature and published charts

There is a published centile chart for Coffin-Siris Syndrome (CSS; OMIM #135900), caused by variants in BAF complex genes (4). Direct comparison with *ARID1B* is not possible given the greater genetic heterogeneity and differing endpoints (height to age 10; OFC to 36 months). In fact, we did not see a sex difference for weight, but the 50^th^ centile for height at age 10, and weight and OFC were consistent.

We were able to analyse the MWS dataset in two ways: LMSz as described here and the conventional LMS method applied to the raw data. The two sets of centiles are reasonably similar though the z-score-based centiles look more convincing for height and weight, particularly at older ages. For BMI, the 2^nd^ and 50^th^ centiles agree well, but the 98^th^ centiles are more discordant. This is probably because the z-score conversion applies the UK90 skewness adjustment to the MWS data, but the degree of skewness in the MWS data is different. With right skewness this would tend to affect the upper rather than the lower centiles of the distribution, as seen here.

MWS centiles have already been published by the MWS Consortium(3), but superficially they look different from the centiles in Figure 4. This may be because we excluded different outliers and fitted a different model–see Supplement 1 for our gamlss code.

There are no published growth charts for *ANKRD11*, *KMT2A*, *DDX3X* or *ASXL3*. Variants in *ANKRD11* cause KBG Syndrome (MIM # 148050) (13). Our growth charts are consistent with the published phenotypes. Growth hormone has been trialled as a therapeutic option for a small number of individuals with short stature, a review of which is underway by Dr Ockeloen’s group (Aukema S. et al, submitted). These growth charts will be an essential tool in monitoring children both in determining need for potential treatment and judging its success.

We selected *DDX3X* syndrome (MIM #300160), a neurodevelopmental disorder predominantly in females (14) to investigate how our charts would work for a very rare gene, where *n*= 5 for the males. Our charts agree with the literature of fairly normal heights and weights in females, with a

proportion having borderline microcephaly. The phenotype in boys is less well understood and the data are limited. Nonetheless our method enables some form of chart to be produced. Considered with caution this could still provide a useful adjunct in a clinical setting, particularly if the data points are viewable in the chart to alert the clinician to the small sample size.

Alterations in *KMT2A* cause Wiedemann-Steiner Syndrome (WSS) (MIM # 605130 (15). WSS is associated with short stature in about 60% of individuals, microcephaly in a third, and weight below the 5^th^ centile in a third. Our charts are concordant with published descriptions depicting median height, weight, and OFC for *KMT2A* at the reference 2^nd^, 2^nd^ and 25^th^ centiles respectively.

*ASXL3*-related disorder (Bainbridge-Ropers Syndrome, MIM # 615485) is associated with normal birth weight but poor postnatal growth due to feeding issues in infancy, which stabilise following feeding intervention (16–18). BMI rises with age which may be explained by sustained feeding intervention or because of dysregulated or impulsive eating behaviours that can develop later in childhood. Feeding interventions should not target reference height (*ASXL3* median height = 12^th^ centile) and our charts will be important for growth monitoring. Median OFC is on the 3^rd^ centile, consistent with published literature of postnatal microcephaly.

Pre- and postnatal growth restriction, sometimes with microcephaly, can be found in up to 34% of individuals with variants in *SATB2* (19, 20) (MIM # 608148). Growth charts were produced recently (1508 data points) but they are hard to compare with ours as they included single variants together with chromosomal microdeletions; and they stopped at age 10 (5). They presented z-score heatmaps which indicate that the microdeletion group was most affected by growth restriction, and this probably explains the difference with our charts.

### Limitations

Most individuals were likely Caucasian although ethnicity was not formally recorded. Data were collated from various sources and some outliers may be due to measurement error, though we sought out extreme measurements and either corrected or excluded them. Fewer OFC measurements were available and birth OFC measurements were frequently missing. UK90 was used as the baseline reference for three reasons: most of the data were from the UK (except for the European ANKRD11 and MOWS and International *ASXL3* cohorts), UK90 is the official UK growth reference for data at birth and over 4 years of age (21), and the DECIPHER database uses UK90 to display the growth data. However, there are inter-country differences in height for any child – see Supplement 2 – and this is true for neurodevelopmental disorders as shown with *ANKRD11 -* clinical interpretation needs to take this into account.

The LMSz method converts raw data to z-scores using the baseline reference, and then models the z-scores. It assumes that any skewness in the raw data matches the skewness in the reference, and this may not be the case. If the two skewness patterns differ it will introduce bias, as seen in Figure 4B for BMI, where the 98^th^ centiles based on the LMS and LMSz methods are very different, particularly in males. BMI is markedly right skew at older ages (i.e. the L value is large and negative), which affects the 98^th^ centile more than the 2^nd^ and 50^th^ centiles.

### Implementation

We plan to automate LMSz within the DECIPHER platform (22, 23) using its open access datasets alongside further growth measurements being collated through the GenROC study (7). Gene-specific growth charts will be viewable within DECIPHER. A future development would be to variant subtype specific charts within our proposed pipeline.

## Conclusions

Children with genetic syndromes often have abnormal growth. LMSz allows clinically useful growth charts to be generated from small datasets. These growth charts are gene specific which are essential for accurate clinical management of children with genetic conditions. We plan to automate our method in DECIPHER to enable these charts to be available in the future for all paediatric disorder genes.

## Data availability

Sequence and variant-level data and phenotypic data for the DDD study data are available from the European Genome-phenome Archive (EGA; https://www.ebi.ac.uk/ega/) with study ID EGAS00001000775. Clinically interpreted variants and associated phenotypes from the DDD study and GenROC study are available through DECIPHER (https://www.deciphergenomics.org/).

## Acknowledgements

We thank all the individuals in the DDD study, GenROC study and *ASXL3* Natural History study and other cohorts who consented for anonymised data sharing. We thank the Mowat Wilson Syndrome Growth Chart Consortium for providing us with their dataset. We thank Lucy Pollock (a member of the GenROC study PPI group) for drawing the original artwork in the graphical abstract.

The authors of this publication are members of the European Reference Network on Rare Congenital Malformations and Rare Intellectual Disability ERN-ITHACA [EU Framework Partnership Agreement ID: 3HP-HP-FPA ERN-01-2016/739516].

The GenROC study would not be possible without the GenROC consortium which is comprised multiple contributors from NHS sites across the UK. The consortium is comprised of individuals who have either acted as a local site PI or Associate PI or completed clinical proformas for GenROC. The list of individuals is as follows: Suzanne Alsters, Ruth Armstrong, Tazeen Ashraf, Queenstone Baker, Diana Baralle, Jonathen Berg, Marta Bertoli, Thomas Boddington, Moira Blyth, Catherine Breen, Helen Brittain, Lisa Bryson, Jenny Carmichael, Emma Clement, Tessa Coupar, Anna de Burca, Cristina Dias, Fleur Dijk, Abhijit Dixit, Alan Donaldson, Andrew Douglas, Jacqueline Eason, Fayadh Fauzi, Elaine Fletcher, Helen V. Firth, Nicola Foulds, Caroline Furnell, Andrew Fry, Laura Furness, Jennifer Gardner, Merrie Gowie, Rachel Harrison, Verity Hartill, Lizzie Harris, Eleanor Hay, Jenny Higgs, Simon Holden, Daniela Iancu, Rachel Irving, Vani Jain, Rosalyn Jewell, Gabriela Jones, Beckie Kaemba, Arveen Kamath, Ayse Nur Kavasoglu, Mira Kharbanda, Sophie King, Alison Kraus, Ajith Kumar, Katherine Lachlan, Neeta Lakhani, Wayne Lam, Anne Lampe, Abigail Lazenbury, Helen Leveret, Jessica Maiden, Alison Male, Alisdair McNeil. Ruth McGowan, Holly McHale, Catherine McWilliam, Jonathan Memish, Lara Menzies, Radwa Mohamed, Tara Montgomery, Oliver Murch, Michael Parker, Caroline Pottinger, Vijayalakshmi Ramakumaran, Ruth Richardson, Alison Ross, Claire Searle, Charles Shaw-Smith, Suresh Somarathi, Edward Steel, Helen Stewart, Kerra Templeton, Riya Tharakan, Madeline Tooley, Mohamed Wafik, Emma Wakeling, Elizabeth Wall, Amy Watford, Patricia Wells, Louise Wilson

## Author contributions

Conceptualization, Investigation, Methodology: KJL, TJC, HVF Formal analysis: TJC Project administration, Supervision, Writing-original draft: KJL, TJC Data curation, Writing-review & editing: all authors.

## Ethics declaration

The DDD study has UK Research Ethics Committee approval (10/H0305/83, granted by the Cambridge South REC, and GEN/284/12 granted by the Republic of Ireland REC). All participants gave informed consent, as required by the REC. All published data were de-identified. Specific ethical approval for the growth charts development was given via a DDD Complementary Analysis Proposal Approval (CAP#371). The GenROC study received East Midlands - Nottingham Research Ethics Committee (REC) approval on 15 December 2022 and Health Research Authority approval on 9 February 2023. The *ASXL3* Natural History Study, sponsored by Sheffield Children’s Hospital and The University of Sheffield (UK) received REC (23/SC/0151) and HRA approval on 2 June 2023. All participants enrolled in the study gave informed consent for anonymised data sharing to allow this collaboration.

