## Supplementary figures and images for "The LMSz method - an automatable scalable approach to constructing gene-specific growth charts in rare disorders"

### Supplementary Figure 2

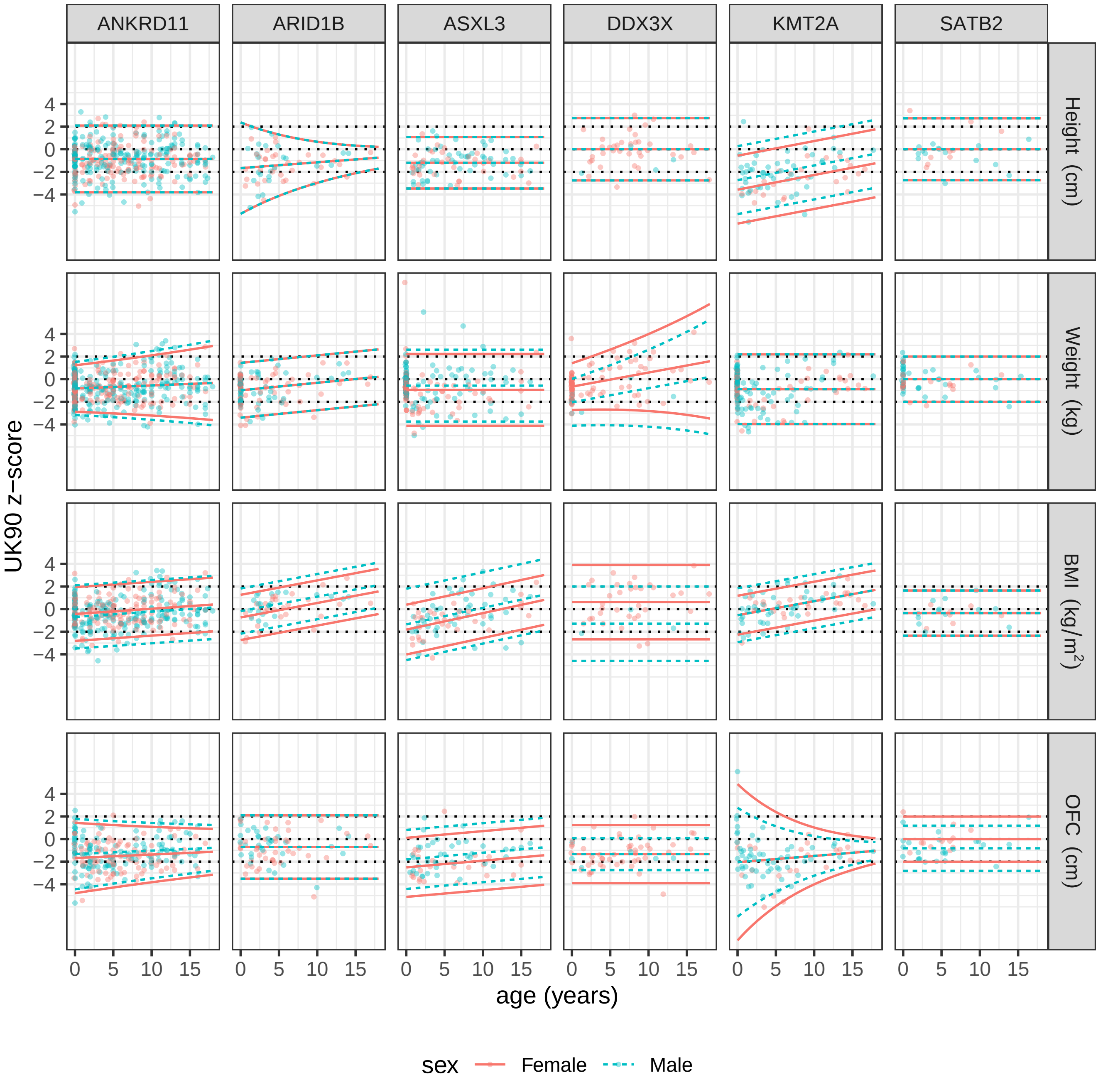

### Supplementary Figure 3

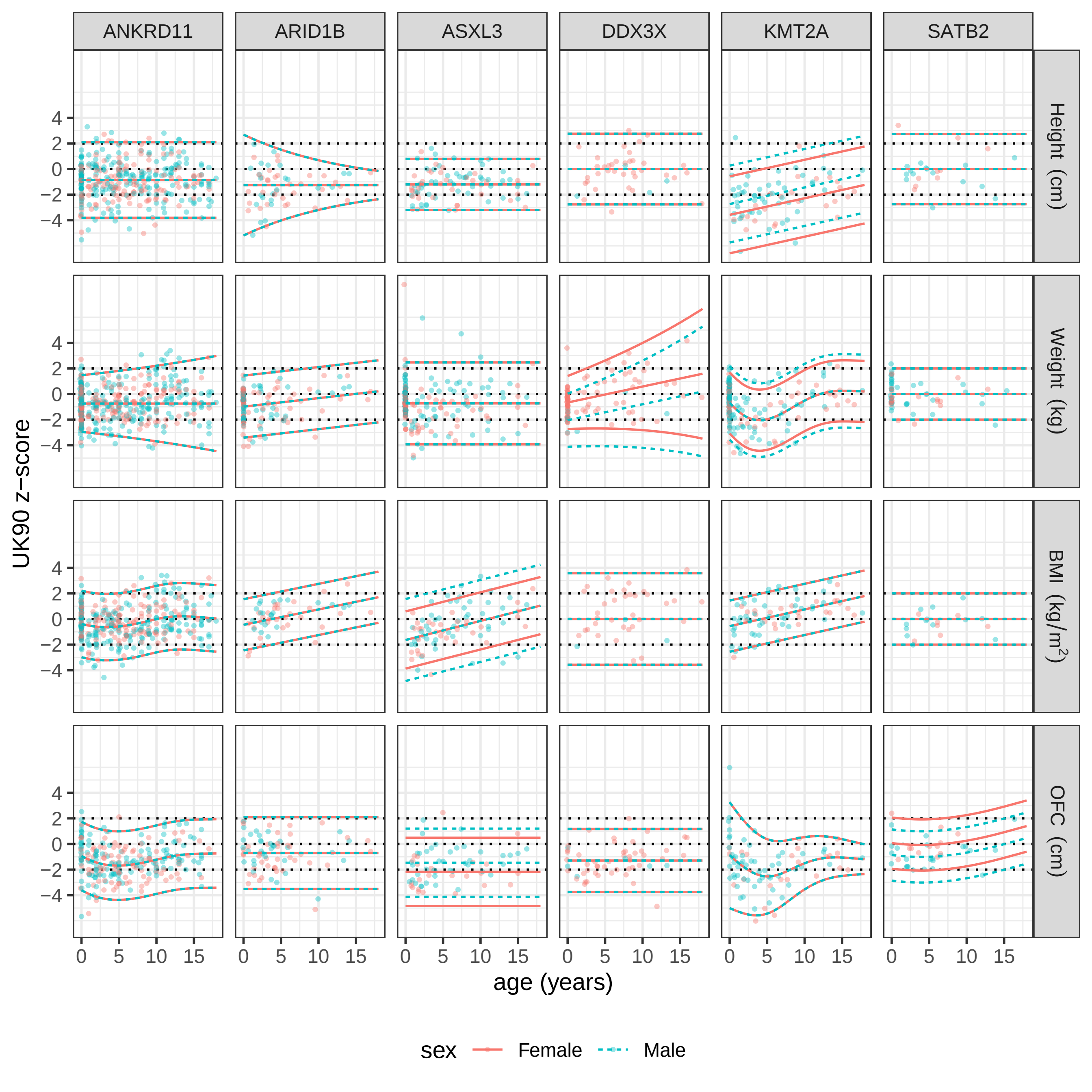

### Supplemetary Figure 1

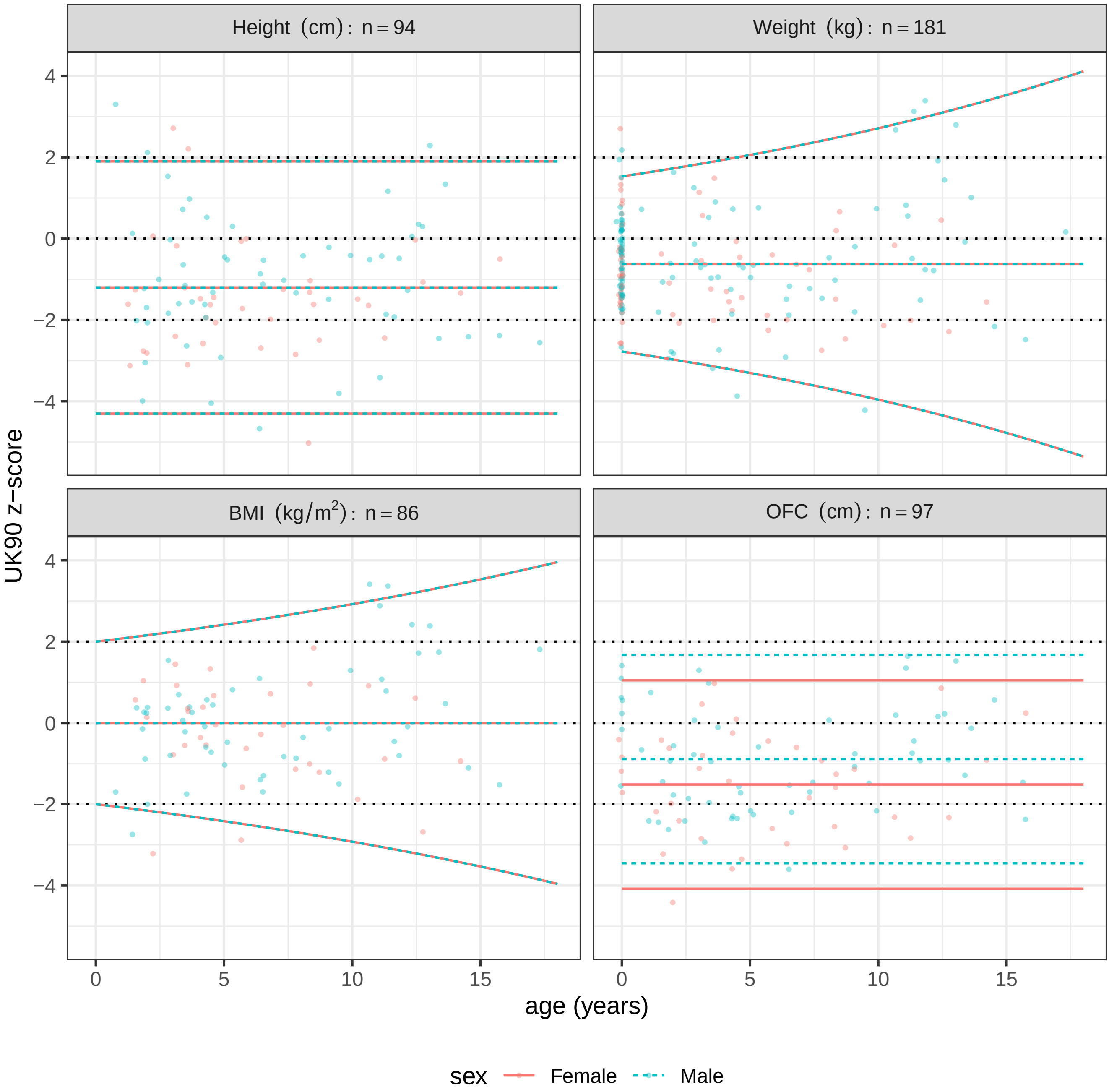
