## Supplementary Table 3 for "The LMSz method - an automatable scalable approach to constructing gene-specific growth charts in rare disorders"

| Measurement | ANKRD11 | ARID1B | ASXL3 | DDX3X | KMT2A | SATB2 |
|---|---|---|---|---|---|---|
| 2nd centile (as z-score) | | | | | | |
| Height | -3.8 | -5.2 | -3.2 | -2.8 | -6.2 | -2.7 |
| Weight | -3 | -3.4 | -3.9 | -3.4 | -4 | -2 |
| BMI | -3.2 | -2.5 | -4.4 | -3.6 | -2.6 | -2 |
| OFC | -4.1 | -3.5 | -4.5 | -3.8 | -6.3 | -2.4 |
| 50th centile | | | | | | |
| Height | 20 | 11 | 12 | 50 | 0.08 | 50 |
| Weight | 23 | 16 | 23 | 8.8 | 19 | 50 |
| BMI | 28 | 33 | 5 | 50 | 29 | 50 |
| OFC | 9.8 | 24 | 3.4 | 9.9 | 5.5 | 34 |
| 98th centile | | | | | | |
| Height | 98 | 100 | 79 | 100 | 44 | 100 |
| Weight | 93 | 93 | 99 | 76 | 99 | 98 |
| BMI | 98 | 94 | 86 | 100 | 93 | 98 |
| OFC | 93 | 98 | 80 | 88 | 100 | 94 |
