## Supplement 1 for "The LMSz method - an automatable scalable approach to constructing gene-specific growth charts in rare disorders"

### Supplement 1. Analysis of Mowat Wilson Syndrome data

We undertook z score scale analysis of respectively 1012, 1106, 999 and 741 measurements for height, weight, BMI and OFC. In addition to the z-score analysis, the MWS raw data were also analysed using the LMS method, due to the relatively large sample size. Prior to analysis, the data were checked for outliers as described in the main text. A total of 37 measurements were excluded, where the internal z-score exceeded 3 in absolute value.

The GAMLSS code to fit the model was as follows:

library(gamlss)

control <- pb.control(inter = 10)

model <- gamlss(value ~ pb((age+3/4)^1/4, max.df = 6, control = control),

sigma.fo = ~pb(age, max.df = 3, control = control),

data = data, family = BCCGo, nu.start = 1,

nu.fix = measure %in% c('Height', 'OFC'))

Here value corresponds to height, weight, BMI or OFC. The median mu curve is fitted as a P-spline in age restricted to 6 or fewer degrees of freedom (d.f.), where age is transformed to over-sample the early measurements – see Cole (2021). The sigma curve is a P-spline with 3 or fewer d.f., and the nu curve is a constant. In addition nu is forced to 1, i.e. a Normal distribution, for height and OFC.

Cole TJ. 2021. Sample size and sample composition for constructing growth reference centiles. Stat Methods Med Res 30:488-507.
