## Supplement 2 for "The LMSz method - an automatable scalable approach to constructing gene-specific growth charts in rare disorders"

Supplement 2. Height differences by country

Figures 1, 2 and 3 show results for height in ANKRD11 based on pooled data from the UK, Spain, Denmark, and the Netherlands. The fitted z-score model consists of intercepts of -0.85 for the mean and 0.39 for the log SD (i.e. SD = 1.5) (see Supplementary Table 1), indicating marked short stature and increased variability compared to the UK90 reference.

To test for the possibility of inter-country differences in mean height, the model was extended to include an adjustment for *country*, and the country mean coefficients are shown in the table, with the overall mean subtracted. Thus the values indicate the country-specific offset to be applied to the centiles from the pooled model in Figure 1. For example centiles for UK children are on average about 0.33 z-scores (half a centile channel width) lower, and Dutch children half a channel width higher, than in Figure 1. These country differences can be taken into account by clinicians when managing their patients.

|  | UK n = 94 | Spain n = 132 | Denmark  n = 26 | Netherlands  n = 103 |
| --- | --- | --- | --- | --- |
| Mean | -0.35 | -0.10 | 0.23 | 0.38 |
| Std. Error | 0.15 | 0.13 | 0.28 | 0.14 |

So Dutch children are highly significantly taller than UK children, by more than a channel width on the chart, which implies that *country* should be added to the regression model. However the penalty associated with adding the three extra degrees of freedom to the model proved to be larger than the reduction in deviance, so its BIC was larger than the simpler model. Hence even if the list of models used in the LMS-z method had included *country*, it would not have been selected.

Non-syndromic children are also known to be taller in the Netherlands than the UK, and the same may well apply to the other syndrome groups in Figure 1. The centiles for the non-ANKRD11 genes are based entirely on UK children, so they may be up to a channel width too low for Dutch children, though this has to be speculative given the absence of further Dutch data.
